# Body Image, Obesity, and Sexual Cohesion: Impacts on Depression among Nigerian University Students

**DOI:** 10.1101/2025.02.02.25321550

**Authors:** Oluwasegun Akinyemi, Olajumoke Kemi Ekundayo, Mojisola Fasokun, Fadeke Ogunyankin, Oghenekaro Samuel Ifoto, Oluwaferanmi Deborah Alatise, Oluebubechukwu Eze, Muyiwa Sunday Okusami, Kakra Hughes, Miriam Michael, Akinola Akinmade

## Abstract

**Introduction:** University students face a variety of challenges, including mental health issues, which are often compounded by societal and individual factors such as body image concerns, obesity, and experiences of intimate partner violence. These factors may adversely affect their mental health and academic performance. Yet, limited research exists on studies evaluating the impact of these factors on depression in Nigerian institutions of higher learning. This study aims to address this gap by examining the impact of these factors on self-reported depression with a focus on the moderating role of sex.

**Objective:** To assess the associations between body image concerns, obesity, intimate partner violence, and sexual cohesion with depression among university students in Nigeria and to explore how these relationships vary by sex.

**Methodology:** This cross-sectional study was conducted over a one-month period among university students in Nigeria. Data was collected through structured, self-administered questionnaires. The primary outcome variables were self-reported depression. Explanatory variables included body image concerns, BMI categories (obese vs. normal BMI), intimate partner violence, and sexual cohesion. Sex was examined as a moderator.

Inverse probability weighting was used to account for confounding variables, including age, sex, year in school, parental education, household income, smoking and alcohol consumption, and other comorbidities. Multivariable regression analyses were performed to evaluate the relationships between explanatory variables and outcomes, adjusting for potential confounders.

**Results:** The study included 501 participants, with 64.5% females and 35.6% males. Most respondents (83.4%) were aged 18–20 years. Obesity was observed in 18.6% of participants, higher in females (20.7%) than males (14.6%).

Sexual coercion was reported by 10.8% (males: 5.6%; females: 13.6%), while 3.4% experienced intimate partner violence (IPV), with similar rates in both genders. Depression was reported by 33.5% of participants, more common in females (35.3%) than males (30.3%).

Body image concerns increased the risk of depression by 35.3% (95% CI: 13.0%-57.7%, p = 0.002), particularly in males (26.3%, 95% CI: 16.4%-69.1%, p = 0.227). Obesity was linked to significantly higher depression rates in males (25.9%, 95% CI: 1.9%-50.0%, p = 0.035) but not in females. Sexual coercion strongly correlated with higher depression rates in both genders (males: 43.0%, 95% CI: 23.5%-62.6%, p < 0.001; females: 39.5%, 95% CI: 20.9%-58.1%, p < 0.001). IPV showed a weaker link to depression, with rates of 21.1% in males and 30.1% in females, though not statistically significant.

**Conclusion:** This study highlights the complex interplay between psychosocial factors and their impact on mental health outcomes among university students in Nigeria. Addressing these factors, particularly through gender-sensitive interventions, is crucial for improving student mental health.

**Policy Implications:** The findings call for the integration of mental health and psychosocial support services in university settings, including counseling and educational programs on body image and intimate partner violence. Policymakers and university administrators should prioritize gender-sensitive approaches to address the unique challenges faced by male and female students. Additionally, strategies to promote healthy lifestyle behaviors and prevent obesity among students should be implemented to enhance their mental health and academic performance.

## INTRODUCTION

Depression is a significant global public health issue and a leading contributor to disability, affecting over 280 million people worldwide (1, 2). Among university students, the prevalence of depression is alarmingly high, with estimates ranging from 10% to 30% globally (3–5). University students are particularly vulnerable due to the transitional nature of their academic and personal lives, which often involves managing academic pressures, social relationships, financial constraints, and future career uncertainties (6–7). In low- and middle-income countries (LMICs), including Nigeria, the burden of depression among university students is particularly pronounced, with studies indicating that approximately one-quarter to one-third of students exhibit depressive symptoms, and some research reporting even higher rates (7–10).

Several psychosocial factors have been identified as significant contributors to depression (11, 12). Body image concerns, defined as negative perceptions, thoughts, and feelings about one’s physical appearance, are particularly associated with low self-esteem, social anxiety, and depressive symptoms, especially in the context of increasing obesity rates (13–15). Obesity, defined by a Body Mass Index (BMI) of 30 or higher, not only carries physical health risks but also contributes to psychological distress due to societal stigma and discrimination (16–18). These challenges are particularly relevant in university settings, where social comparison and peer influence are prevalent.

Intimate partner violence (IPV), encompassing physical, emotional, and sexual abuse within a romantic relationship, is another critical factor affecting students’ mental health and academic outcomes (19, 20). IPV is associated with a heightened risk of depression, post-traumatic stress disorder, and disruptions in daily functioning. In university populations, the prevalence of IPV is substantial, and its psychological and academic consequences are profound (20, 21).

Sexual cohesion, referring to mutual understanding and harmony in sexual relationships, is a less explored variable but may play a significant role in influencing mental health (22, 23). Inconsistent or negative experiences in sexual relationships can exacerbate feelings of isolation, distress, and depression, further impacting academic outcomes (24, 25). Exploring the interplay between sexual cohesion and other psychosocial factors is essential, particularly in culturally diverse settings like Nigeria, where societal norms and gender dynamics shape relationship experiences.

While these factors affect a lot of students, their impact may vary significantly between genders. Sex differences are a crucial consideration in understanding the relationships between these psychosocial factors, depression, and academic performance (26, 27). Evidence suggests that female students may be more vulnerable to body image concerns and IPV, leading to higher rates of depression and academic difficulties compared to their male counterparts (28, 29). Understanding these gender-specific dynamics is essential for developing targeted interventions.

Despite the growing recognition of these issues, research on the combined impact of body image concerns, obesity, IPV, and sexual cohesion on depression in Nigerian university students remains limited. Additionally, the moderating role of sex in these relationships is underexplored. Addressing these gaps is critical for informing gender-sensitive policies and interventions aimed at improving mental health outcomes among university students.

This study aims to investigate the associations between body image concerns, obesity, IPV, and sexual cohesion with self-reported depression, with a particular focus on the moderating role of sex. The findings are expected to offer valuable insights into the psychosocial determinants of mental health, contributing to the development of effective strategies for promoting student well-being in Nigerian universities.

## METHODOLOGY

### Study Design

This study utilized a cross-sectional design to examine the associations between body image concerns, obesity, IPV, sexual cohesion, and depression among university students in Nigeria. The moderating role of sex in these relationships was also investigated.

### Study Setting and Population

The study was conducted over a one-month period from September 20, 2024, to October 21, 2024, at Afe Babalola University, a private university in Nigeria. Participants were undergraduate students aged 18 years or older enrolled in various academic programs. A diverse sample was recruited to ensure representation across different academic disciplines, years of study, and socioeconomic backgrounds. Written informed consent was obtained from all participants before data collection, and a questionnaire was distributed to gather relevant study data.

### Sample Size and Sampling Technique

Based on power calculations, a sample size of 500 students was determined to detect moderate effect sizes for the primary outcomes. Participants were selected using stratified random sampling, with strata defined by academic year and faculty. This approach ensured a proportional representation of students from different academic levels and disciplines.

The sample size for this study was calculated to determine the minimum number of participants needed to detect statistically significant associations between selected factors and depression among university students in Nigeria. Based on prior research, the prevalence of depression was estimated at 25% (10). Using a confidence level of 95% (Z = 1.96) and a margin of error of 5% (e = 0.05), the initial sample size was computed using the formula:

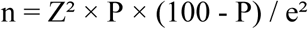

This calculation yielded approximately 288 participants. To account for potential non-response, the sample size was adjusted by adding 10%, resulting in a final required sample size of 317 participants. However, at the conclusion of the study, data were collected from 501 students, exceeding the calculated sample size. All respondents were included in the final analysis.

Participants were selected using stratified random sampling, with strata defined by academic year and faculty to ensure proportional representation across different academic levels and disciplines.

### Data Collection

Data was collected using a structured, self-administered questionnaire. The questionnaire consisted of validated scales to measure the primary outcomes and explanatory variables:

- **Depression:** Assessed using the Patient Health Questionnaire-9 (PHQ-9), a widely used tool for measuring depressive symptoms (30).
- **Body Image Concerns:** Evaluated using the Body Image Satisfaction Scale (BISS) (31, 32).
- **Obesity:** Classified based on Body Mass Index (BMI) categories derived from self-reported height and weight (33, 34).
- **Intimate Partner Violence (IPV):** Assessed using the Conflict Tactics Scale (CTS2) (35).
- **Sexual Cohesion:** Measured using items adapted from validated relationship cohesion scales (36).

### Confounding Variables

The following variables were included as potential confounders in the analysis:

- **Demographic factors:** Age, sex, year in school, parental education, and household income.
- **Lifestyle behaviors:** Smoking, alcohol consumption, and physical activity.
- **Health factors:** Presence of comorbidities.

### Data Analysis

Data was analyzed using Stata software. Descriptive statistics were used to summarize participant characteristics and study variables. Bivariate analyses were conducted to explore relationships between explanatory variables and outcomes.

Inverse probability weighting (IPW) was employed to adjust for confounding variables. Multivariable regression models were constructed to assess the independent associations between body image concerns, obesity, IPV, and sexual cohesion with depression. Interaction terms were included to evaluate the moderating effect of sex.

### Ethical Considerations

The study received ethical approval from Afe Babalola University’s Institutional Review Board (AMSH/REC/24/089). Prior to data collection, all participants provided written informed consent. Participation was voluntary, and confidentiality was ensured throughout the study, with personal identifiers removed from the dataset.

## RESULT

### Baseline Characteristics

Table 1 highlights the baseline characteristics of the study respondents, stratified by sex. The study population comprised 501 university students, with 178 males (35.5%) and 323 females (64.5%). Most participants were between 18–20 years of age (83.4%), with no significant difference between males (85.4%) and females (82.4%). Regarding maternal educational attainment, 33.3% of the participant’s mothers had advanced education, while 42.9% had tertiary education, and 23.8% reported high school or lower education. Household income was predominantly greater than 150,000 NGN for 78.8% of the participants, and no significant differences were observed between males and females (p=0.445). Most participants came from monogamous family structures (83.2%), with fewer from polygamous (9.6%) or single-parent households (7.2%), with no significant differences between genders (p=0.788).

**Table 1:** Baseline Characteristics of Study Population.

| Variables | Total Population (N=501) | Male (n=178) | Female (n=323) | Chi-Squared | p-value |
| --- | --- | --- | --- | --- | --- |
| **Age** |  |  |  | 2.1154 | 0.347 |
| **18-20Yr.** | 418 (83.4%) | 152 (85.4%) | 266 (82.4%) |  |  |
| **21-24Yr.** | 75 (15.0%) | 22 (12.4%) | 53 (16.4%) |  |  |
| **>24Yr.** | 8 (1.6%) | 4 (2.3%) | 4 (1.2%) |  |  |
| **Maternal Education** |  |  |  | 10.549 | 0.005 |
| **High School or Lower** | 119 (23.8%) | 54 (30.3%) | 65 (20.1%) |  |  |
| **Tertiary** | 215 (42.9%) | 79 (44.4%) | 136 (42.1%) |  |  |
| **Advanced** | 167 (33.3%) | 45 (25.3%) | 122 (37.8%) |  |  |
| **Household Income** |  |  |  | 0.5825 | 0.445 |
| **<=150K** | 106 (21.2%) | 41 (23.0%) | 65 (20.0%) |  |  |
| **>150K** | 395 (78.8%) | 137 (77.0%) | 258 (79.9%) |  |  |
| **Family Structure** |  |  |  | 0.4776 | 0.788 |
| **Monogamous** | 417 (83.2%) | 149 (83.7%) | 268 (83.0%) |  |  |
| **Polygamous** | 48 (9.6%) | 18 (10.1%) | 30 (9.3%) |  |  |
| **Single Parent** | 36 (7.2%) | 11 (6.2%) | 25 (7.7%) |  |  |
| **Explanatory Variables** |  |  |  |  |  |
| **IPV** | 17 (3.4%) | 6 (3.4%) | 11 (3.4%) | 0.0004 | 0.984 |
| **Coerced Sex** | 54 (10.8%) | 10 (5.6%) | 44 (13.6%) | 7.6455 | 0.006 |
| **BMI** |  |  |  | 8.1824 | 0.042 |
| **Normal weight** | 245 (48.9%) | 102 (57.3%) | 143 (44.3%) |  |  |
| **Underweight** | 53 (10.6%) | 15 (8.4%) | 38 (11.8%) |  |  |
| **Overweight** | 110 (22.0%) | 35 (19.7%) | 75 (23.2%) |  |  |
| **Obese** | 93 (18.6%) | 26 (14.6%) | 67 (20.7%) |  |  |
| **Lifestyle Behaviors** |  |  |  |  |  |
| **Alcohol** | 104 (20.8%) | 39 (21.9%) | 65 (20.1%) | 0.2226 | 0.637 |
| **Smoking** | 22 (4.4%) | 8 (4.5%) | 14 (4.3%) | 0.007 | 0.933 |
| **Outcomes** |  |  |  |  |  |
| **Depression** | 168 (33.5%) | 54 (30.3%) | 114 (35.3%) | 1.2652 | 0.261 |
| **CGPA** | 4.1 ±0.6 | 4.0 ± 0.7 | 4.2 ±0.6 | t=-3.499 | 0.001 |

In terms of explanatory variables, 3.4% of students reported experiencing intimate partner violence (IPV), with no difference between males and females (p=0.984). However, coerced sex was significantly more prevalent among females (13.6%) than males (5.6%) (p=0.006). BMI categories revealed that 48.9% of participants were of normal weight, 22.0% overweight, 18.6% obese, and 10.6% underweight. Obesity was more common among females (20.7%) than males (14.6%) (p=0.042). For lifestyle behaviors, alcohol consumption was reported by 20.8% of participants, while 4.4% reported smoking, with no significant differences by gender for either behavior.

33.5% of participants had experienced some form of depression, with a higher prevalence among females (35.3%) compared to males (30.3%), though this difference was not statistically significant (p=0.261). Academic performance, measured by CGPA, was higher among females (4.2 ± 0.6) compared to males (4.0 ± 0.7) (p=0.001) (Table 1).

### Predicting Depression among Male Students by Body Mass Index (BMI)

Table 2 presents the Average Treatment Effect of BMI categories on the incidence of depression among male respondents. The relationship between the BMI categories and depression among male students was analyzed using an inverse probability weighting approach. Students with a normal BMI had a depression incidence rate of 17.0% (Coef. = 0.1701, 95% CI: 0.0875–0.2527, p<0.001).

**Table 2:** Average Treatment Effect of BMI Categories on Depression Incidence (Male) shows the Average Treatment Effects (ATE) of BMI categories on depression incidence among male university students in Nigeria, using inverse probability weighting (IPW) to adjust for confounders like age and lifestyle factors. Depression risk is compared across BMI groups (Underweight, Overweight, Obese) relative to the Normal BMI group (reference). The table provides coefficients (Coef.), standard errors (Std. Err.), z-scores (z), p-values (P>z), and 95% confidence intervals (95% CI). Overweight and Obese students showed significantly higher depression risks, while no significant effect was observed for the Underweight grou*p*.

| Outcome | BMI (Treated vs. Control) | Coef. | Std. Err. | z | P>z | 95% CI |  |
| --- | --- | --- | --- | --- | --- | --- | --- |
| Depression | Underweight vs. Normal BMI | -0.012 | 0.077 | -0.150 | 0.881 | -0.162 | 0.139 |
|  | Overweight vs. Normal BMI | 0.196 | 0.100 | 1.960 | 0.050 | 0.000 | 0.391 |
|  | Obese vs. Normal BMI | 0.259 | 0.123 | 2.110 | 0.035 | 0.019 | 0.500 |
|  | Normal (reference) | 0.170 | 0.042 | 4.040 | 0.000 | 0.088 | 0.253 |

Overweight students experienced a 19.6% higher incidence of depression compared to those with a normal BMI (Coef. = 0.1959, 95% CI: 0.0004–0.3913, p=0.05). Similarly, obese students showed a significant positive association with depression, further increasing the odds of depressive symptoms by 25.9% (Coef. = 0.2594, 95% CI: 0.0187–0.5001, p=0.035). However, underweight students showed no significant association with depression (Coef. = -0.0115, 95% CI: -0.1615 to 0.1385, p=0.881) (Table 2).

### Predicting Depression among Female Students by BMI

Table 3 presents the Average Treatment Effect of BMI categories on the incidence of depression among female respondents. Female students with normal BMI reported the highest incidence of depression among students with a normal BMI (Coef. = 0.3999, 95% CI: 0.3219–0.4779, p<0.001).

**Table 3:** Average Treatment Effect of BMI Categories on Depression Incidence (Female) presents the Average Treatment Effects (ATE) of BMI categories on depression incidence among female university students in Nigeria, using inverse probability weighting (IPW) to adjust for confounders. Depression risk is compared across BMI categories (Underweight, Overweight, Obese) relative to the Normal BMI group (reference). The table provides coefficients (Coef.), standard errors (Std. Err.), z-scores (z), p-values (P>z), and 95% confidence intervals (95% CI). Overweight students showed a significant reduction in depression risk compared to those with Normal BMI, while no significant effects were observed for the Underweight and Obese groups. Students with Normal BMI had a significant baseline risk of depression.

| Outcome | BMI (Treated vs. Control) | Coef. | Std. Err. | z | P>z | 95% CI |  |
| --- | --- | --- | --- | --- | --- | --- | --- |
|  | Underweight vs. Normal BMI | -0.121 | 0.084 | -1.450 | 0.146 | -0.285 | 0.042 |
|  | Overweight vs. Normal BMI | -0.141 | 0.061 | -2.290 | 0.022 | -0.261 | -0.020 |
|  | Obese vs. Normal BMI | 0.017 | 0.068 | 0.250 | 0.803 | -0.117 | 0.151 |
|  | Normal (reference) | 0.400 | 0.040 | 10.040 | 0.000 | 0.322 | 0.478 |

Overweight students demonstrated a negative association with depression, suggesting a lower likelihood of depressive symptoms compared to those with a normal BMI (Coef. = -0.1406, 95% CI: -0.2611 to -0.0201, p=0.022). However, underweight students showed no significant difference from the baseline, which were students with a normal BMI (Coef. = -0.1213, 95% CI: -0.2851 to 0.0424, p=0.146). Similarly, obesity was not significantly associated with depression in this cohort (Coef. = 0.0170, 95% CI: -0.1167 to 0.1508, p=0.803) (Table 3).

### Depression among Male Students across Body Image Concerns

Table 4 illustrates the impact of body image concerns on the incidence of depression among male student respondents. Students with moderate body image concerns had a significantly higher likelihood of reporting depression compared to those with low concerns (Coef. = 0.4207, 95% CI: 0.1966–0.6447, p<0.001). Though students with high body image concerns have a 26.3% higher incidence of depression than the baseline, this was not statistically significant depression (Coef. = 0.2633, 95% CI: -0.1640–0.6906, p=0.227). Students with low body image concerns have a 26.2% adjusted incidence of depression (Coef. = 0.2621, 95% CI: 0.1901– 0.3342, p<0.001) (Table 4).

**Table 4:** Average Treatment Effect of Body Concerns on Depression Incidence (Male) presents the Average Treatment Effects (ATE) of body image concerns on depression incidence among male university students in Nigeria, using inverse probability weighting (IPW) to adjust for confounders. Depression risk is compared between students with Moderate and High body image concerns relative to those with Low concerns (reference group). The table includes coefficients (Coef.), standard errors (Std. Err.), z-scores (z), p-values (P>z), and 95% confidence intervals (95% CI). Moderate body image concerns were significantly associated with a higher risk of depression, while no significant effect was observed for High concerns compared to the Low concern group.

| Outcome | Body Image (Treated vs. Control) | Coef. | Std. Err. | z | P>z | 95% CI |  |
| --- | --- | --- | --- | --- | --- | --- | --- |
| Depression | Moderate Concern vs. Low | 0.421 | 0.114 | 3.680 | 0.000 | 0.197 | 0.645 |
|  | High Concern vs. Low | 0.263 | 0.218 | 1.210 | 0.227 | -0.164 | 0.691 |
|  | Low Concern (reference) | 0.262 | 0.037 | 7.130 | 0.000 | 0.190 | 0.334 |

### Depression among Female Students across Body Image Concerns

Table 5 illustrates the impact of body image concerns on the incidence of depression among female student respondents. Female students with moderate body image concerns had a 23.7% incidence of depression compared to those with low concerns (Coef. = 0.2370, 95% CI: 0.0983– 0.3757, p=0.001). Similarly, high body image concerns were significantly associated with depression, with an even greater magnitude of effect (Coef. = 0.3532, 95% CI: 0.1296–0.5769, p=0.002).

**Table 5:** Average Treatment Effect of Body Image Concerns on Depression Incidence (Female) presents the Average Treatment Effects (ATE) of body image concerns on depression incidence among female university students in Nigeria, using inverse probability weighting (IPW) to adjust for confounders. Depression risk is compared between students with Moderate and High body image concerns relative to those with Low concerns (reference group). The table includes coefficients (Coef.), standard errors (Std. Err.), z-scores (z), p-values (P>z), and 95% confidence intervals (95% CI). Both Moderate and High body image concerns were significantly associated with an increased risk of depression, with High concerns showing a greater effect. Students with Low concerns had a significant baseline risk of depression.

| Outcomes | Body Image (Treated vs. Control) | Coef. | Std. Err. | z | P>z | 95% CI |  |
| --- | --- | --- | --- | --- | --- | --- | --- |
|  | Moderate Concern vs. Low | 0.237 | 0.071 | 3.350 | 0.001 | 0.098 | 0.376 |
|  | High Concern vs. Low | 0.353 | 0.114 | 3.100 | 0.002 | 0.130 | 0.577 |
|  | Low Concern (reference) | 0.264 | 0.030 | 8.670 | 0.000 | 0.205 | 0.324 |

Students with low body image concerns also (the reference group) reported an adjusted incidence of depression of 26.4% (Coef. = 0.2642, 95% CI: 0.2045–0.3239, p<0.001) (Table 5).

### IPV and Depression among Male Students

Table 6 presents the effect of IPV on the incidence of depression among male student respondents. Male students who did not experience IPV had an adjusted incidence of 30.1% (Coef. = 0.3009, 95% CI: 0.2324–0.3695, p<0.001).

**Table 6:** Average Treatment Effect of IPV on Depression Incidence (Male) shows the Average Treatment Effects (ATE) of intimate partner violence (IPV) on depression incidence among male university students in Nigeria, using inverse probability weighting (IPW) to adjust for confounders. Depression risk is compared between students who experienced IPV and those who did not (reference group). The table provides coefficients (Coef.), standard errors (Std. Err.), z-scores (z), p-values (P>z), and 95% confidence intervals (95% CI). No significant association was found between IPV and depression among those who experienced IPV, while students who did not experience IPV had a significant baseline risk of depression.

| Outcome | IPV (Treated vs. Control) | Coef. | Std. Err. | z | P>z | 95% CI |  |
| --- | --- | --- | --- | --- | --- | --- | --- |
| Depression | Yes | 0.211 | 0.318 | 0.660 | 0.508 | -0.413 | 0.834 |
|  | No (reference) | 0.301 | 0.035 | 8.600 | 0.000 | 0.232 | 0.370 |

Interestingly, male students who experienced IPV had a 21.1% higher adjusted incidence of depression, though the association was not statistically significant (Coef. = 0.2107, 95% CI: - 0.4130 to 0.8344, p=0.508), indicating that IPV may not be a critical determinant of depression in this cohort (Table 6).

### IPV and Depression among Male Students

Table 7 presents the effect of IPV on the incidence of depression among female student respondents. Female students who did not experience IPV had a high incidence of depression (Coef. = 0.3477, 95% CI: 0.2951–0.4003, p<0.001).

**Table 7:**
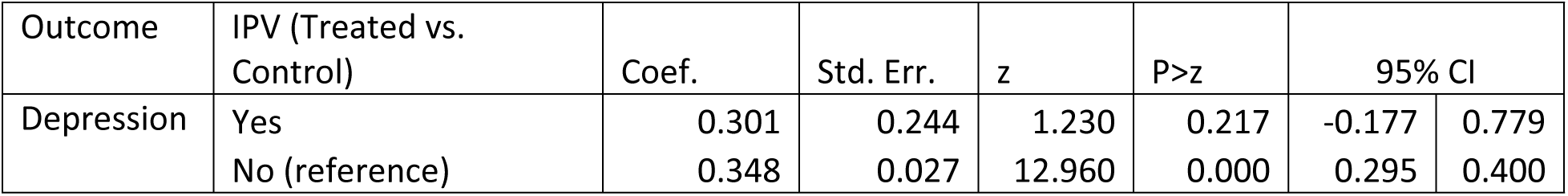
Average Treatment Effect of IPV on Depression Incidence (Female) presents the Average Treatment Effects (ATE) of intimate partner violence (IPV) on depression incidence among female university students in Nigeria, using inverse probability weighting (IPW) to control for confounders. Depression risk is compared between students who experienced IPV and those who did not (reference group). The table provides coefficients (Coef.), standard errors (Std. Err.), z-scores (z), p-values (P>z), and 95% confidence intervals (95% CI). No significant association was observed between IPV and depression among students who experienced IPV, while those who did not experience IPV had a significant baseline risk of depression.

In contrast, for those who experienced IPV, the association with depression was not significantly different from those who did not experience IPV (Coef. = 0.3011, 95% CI: -0.1769 to 0.7792, p=0.217) (Table 7).

### Depression among Male Students by Sexual Cohesion (Coerced Sex)

Table 8 highlights the impact of sexual cohesion on the incidence of depression among male student respondents. Male students who reported experiencing coerced sex had a significantly higher likelihood of reporting depression (Coef. = 0.4304, 95% CI: 0.2346–0.6263, p<0.001). However, those who did not experience coerced sex had an adjusted incidence rate of 29.1% (Coef. = 0.2912, 95% CI: 0.2222–0.3601, p<0.001) (Table 8).

**Table 8:** Average Treatment Effect of Sexual Cohesion on Depression Incidence (Male) presents the Average Treatment Effects (ATE) of sexual cohesion on depression incidence among male university students in Nigeria, using inverse probability weighting (IPW) to control for confounders. Depression risk is compared between students who experienced sexual cohesion and those who did not (reference group). The table includes coefficients (Coef.), standard errors (Std. Err.), z-scores (z), p-values (P>z), and 95% confidence intervals (95% CI). Experiencing sexual cohesion was significantly associated with a higher risk of depression, while students who did not experience sexual cohesion also demonstrated a significant baseline risk of depression.

| Outcome | Cohesion (Treated vs. Control) | Coef. | Std. Err. | z | P>z | 95% CI |  |
| --- | --- | --- | --- | --- | --- | --- | --- |
| Depression | Yes | 0.430 | 0.100 | 4.310 | 0.000 | 0.235 | 0.626 |
|  | No (reference) | 0.291 | 0.035 | 8.280 | 0.000 | 0.222 | 0.360 |

### Depression among Female Students by Sexual Cohesion (Coerced Sex)

Table 9 shows the impact of sexual cohesion on the incidence of depression among female student respondents. Female students who reported experiencing coerced sex had a significantly higher incidence of depression, 39.5% (Coef. = 0.3952, 95% CI: 0.2089–0.5814, p<0.001).

**Table 9:**
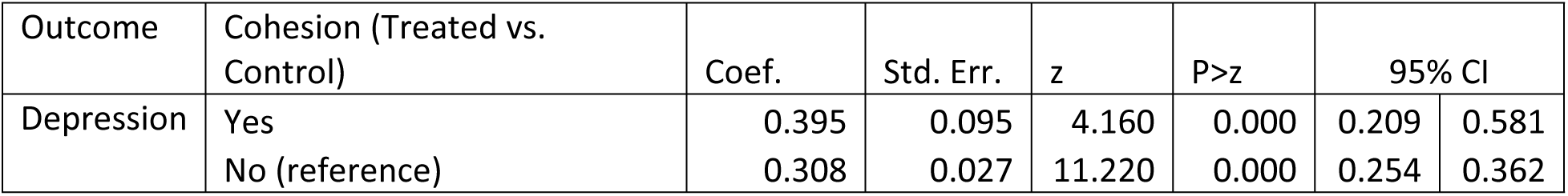
Average Treatment Effect of Sexual Cohesion on Depression Incidence (Female) presents the Average Treatment Effects (ATE) of sexual cohesion on depression incidence among female university students in Nigeria, using inverse probability weighting (IPW) to adjust for confounders. Depression risk is compared between students who experienced sexual cohesion and those who did not (reference group). The table includes coefficients (Coef.), standard errors (Std. Err.), z-scores (z), p-values (P>z), and 95% confidence intervals (95% CI). Experiencing sexual cohesion was significantly associated with a higher risk of depression, while students who did not experience sexual cohesion also had a significant baseline risk of depression.

Female students without such experience reported an adjusted incidence of depression at 30.8% (Coef. = 0.3081, 95% CI: 0.2543–0.3619, p<0.001). The magnitude of this association was smaller compared to those who reported coerced sex.

## DISCUSSION

This study highlights the factors associated with self-reported depression among university students in Nigeria, with rates notably higher among female students than their male counterparts. This finding aligns with global trends, which often show that women are more likely to experience depression due to biological, social, and cultural factors (37–39). However, the high prevalence observed in this population underscores the urgent need for targeted mental health interventions within university settings to address gender-specific vulnerabilities.

One of the most striking findings of this study is the presence of IPV and sexual cohesion issues among male students, challenging traditional beliefs that such exposures predominantly affect women. While IPV and sexual cohesion were reported more frequently among female students, their occurrence among male students suggests a broader societal issue that warrants further investigation highlighting the need for inclusive support systems and education programs (40) that address IPV and sexual coercion across all genders, promoting healthier relationships and reducing the associated mental health burden (41).

Interestingly, no significant association was found between obesity and self-reported depression, particularly among female university students, although the results suggested a possible trend. The lack of statistical significance may be attributed to insufficient sample size or the complex interplay of factors that mediate the relationship between obesity and mental health. These findings align with some previous studies that suggest the relationship between obesity and depression may be influenced by confounding variables, such as stigma, self-esteem, and cultural perceptions of body image (42–45). Further research is needed to explore these dynamics in greater detail.

Body image concerns emerged as a key predictor of self-reported depression, with a high proportion of students expressing moderate to high concerns about their body image. These concerns were more prevalent among female students, reflecting societal pressures and gender norms disproportionately affecting women (28, 45). Similarly, experiencing sexual cohesion was significantly associated with depression, with individuals exposed to these factors experiencing higher odds of depressive symptoms. Although IPV did not demonstrate a significant association, the effect sizes were substantial enough to justify further investigation. These findings underscore the critical role of psychosocial factors in shaping mental health outcomes (20, 46) and highlight the importance of addressing these issues as part of university mental health initiatives.

The findings also emphasize the importance of gender-sensitive interventions. Female students were disproportionately affected by IPV, sexual coercion, and body image concerns, which contributed significantly to their higher rates of depression. Male students, while reporting lower rates of these exposures, also demonstrated significant associations with depression, particularly in the context of sexual coercion. This highlights the need for tailored mental health programs that account for the unique experiences of both genders.

### Strengths and Limitations

#### Strengths

This study has several notable strengths. First, it provides valuable insights into the psychosocial factors influencing depression and academic performance among university students in Nigeria, a context where limited research exists on this topic. By incorporating variables such as body image concerns, intimate partner violence (IPV), sexual cohesion, and obesity, the study adopts a comprehensive approach to understanding the complex interplay of factors affecting mental health and academic outcomes.

Second, the use of a cross-sectional design with a relatively large and diverse sample (N=501) enhances the representativeness of the findings for the studied university population. The stratified random sampling method ensured representation across different academic disciplines, socioeconomic backgrounds, and genders, thereby minimizing selection bias.

Third, the study’s robust analytical approach using inverse probability weighting (IPW) strengthens the validity of the findings. By adjusting for potential confounding variables, such as age, parental education, household income, and lifestyle behaviors, the study isolates the independent effects of explanatory variables on depression and academic performance. This methodology is particularly effective in reducing bias and ensuring reliable estimates of associations.

Lastly, the study’s focus on gender-specific analyses provides nuanced insights into how psychosocial factors impact male and female students differently. This approach underscores the importance of gender-sensitive mental health interventions and contributes to a growing body of literature advocating for tailored mental health strategies.

#### Limitations

Despite its strengths, this study has some limitations. It’s important to note that the study’s cross-sectional nature limits causal inferences and may not fully account for potential bidirectional relationships between variables. While these findings provide valuable insights into the associations between the factors studied in this population, further longitudinal studies would be necessary to examine the temporal relationships and causal pathways underlying these associations

Second, the reliance on self-reported measures for key variables, such as depression, body image concerns, IPV, and sexual cohesion, introduces the possibility of social desirability and recall biases. Participants may have underreported or misreported sensitive information, potentially leading to an underestimation of the prevalence of IPV and sexual coercion.

Third, the study’s focus on a single university may limit the generalizability of the findings to other educational institutions in Nigeria or beyond. Different universities may have varying cultural, social, and environmental contexts that influence the prevalence of depression and associated factors.

Fourth, the absence of detailed clinical data, such as prior mental health diagnoses, access to healthcare services, and comorbidities, constrains the study’s ability to account for these critical factors in the analysis. Similarly, while the study adjusted for confounders such as parental education and income, unmeasured variables, such as social support networks or resilience, may also play significant roles in determining mental health outcomes.

Finally, while obesity and depression were explored, the lack of significant findings could be attributed to insufficient statistical power or the complexity of the relationship. Future studies should use larger sample sizes and more granular measures of obesity, such as waist-to-hip ratio or body fat percentage, to better understand this association.

### Conclusion

The present study provides valuable insights into the complex interplay of psychosocial factors and their impact on depression among university students in Nigeria. The findings highlight the need for comprehensive, gender-sensitive mental health strategies that address body image concerns, IPV, and sexual cohesion. Universities should prioritize creating safe, supportive environments and provide accessible mental health services to mitigate these risk factors and promote the well-being and academic success of their students. Future research should explore longitudinal trends and the effectiveness of targeted interventions in reducing depression and its associated burdens.

## Data Availability

All relevant data are within the manuscript and its Supporting Information files.

